# Early visual processing as a marker of disease, not vulnerability: Event-related potential (ERP) evidence from 22q11.2 deletion syndrome, a population at high risk for schizophrenia

**DOI:** 10.1101/2021.10.26.21265538

**Authors:** Ana A. Francisco, John J. Foxe, Douwe J. Horsthuis, Sophie Molholm

## Abstract

We investigated visual processing in 22q11.2 deletion syndrome (22q11.2DS), a condition characterized by an increased risk for schizophrenia. Visual processing differences have been described in schizophrenia but remain understudied early in the disease course. Electrophysiology was recorded during a visual adaptation task with different interstimulus intervals to investigate visual processing and adaptation in 22q11.2DS (with (22q+) and without (22q-) psychotic symptoms), compared to control and idiopathic schizophrenia groups. Analyses focused on early windows of visual processing. While increased amplitudes were observed in 22q11.2DS in an earlier time window (90-140 ms), decreased responses were seen later (165-205 ms) in schizophrenia and 22q+. 22q11.2DS, and particularly 22q-, presented increased adaptation effects. We argue that while amplitude and adaptation in the earlier time window may reflect specific neurogenetic aspects associated with a deletion in chromosome 22, amplitude in the later window may be a marker of the presence of psychosis and/or of its chronicity/severity.

## Introduction

22q11.2 deletion syndrome (22q11.2DS) is a rare genetic condition characterized by a markedly increased risk for schizophrenia, a severe and often chronic psychiatric disorder. Approximately half of individuals with a chromosome 22 deletion present schizotypical traits and experience transient psychotic states (1) and about 30% receive a formal diagnosis of schizophrenia (2, 3). The study of 22q11.2DS, particularly of the younger individuals with the deletion who have not yet experienced psychotic symptoms or been on antipsychotic medication for a prolonged period, may prove invaluable to the identification of early markers associated with schizophrenia.

Sensory processing differences in the visual domain are commonly reported in the schizophrenia literature (for a review of findings, see (4)). Orientation (5), motion (6-10), contrast sensitivity (11-14), perceptual organization (15-18), spatial discrimination (19-21), detection of targets masked after short intervals (22-25), visual object processing (26), and early visual processing and adaptation (27-34) have all been shown to be impaired in this population. Such sensory impairments may presage higher-order cognitive (35-39) and social (40) deficits and daily functioning difficulties (11, 41), which in turn are related to clinical outcome (42). While early identification of risk and treatment of schizophrenia are key to improving prognosis (43-46), sensory processing remains relatively understudied early in the disease. Indeed, a significant portion of the studies exploring perception and cognition in schizophrenia have been conducted with chronically ill individuals. Though the characterization of sensory/perceptual function in chronic stages of the disease is highly relevant, the effects of prolonged use of antipsychotic medication and continuous hospitalizations may complicate the interpretation of findings. The handful of studies on visual processing and visuo-spatial attention in first-episode and prodromal schizophrenia (38, 47-51) suggest that visual processing deficits are not only seen in chronic schizophrenia, but, rather, are present from early on in the disease course. Furthermore, visual processing deficits have been found in first degree relatives of individuals with schizophrenia (28, 30). Thus, visual processing may hold potential as an endophenotypic marker of genetic risk for developing the condition.

22q11.2DS presents a unique opportunity to extend the study of altered visual processing to a population genetically predisposed to schizophrenia, potentially amplifying effect sizes for the identification of systems involved in psychosis and informing a possible genetic pathway to the disease. To our knowledge, only one behavioral (52) and two electrophysiological (EEG) (53, 54) studies have investigated basic visual processing in 22q11.2DS. While these support the presence of visual processing differences in 22q11.2DS that, descriptively, partially recapitulate those seen in schizophrenia, no differentiation was made between those with the deletion and psychotic symptoms and those with the deletion and no psychotic symptoms. The importance of this distinction is apparent from our previous work (55, 56), in which we demonstrated significant differences in both cognitive and neural function between these two groups. Moreover, the focus on this population and subsequent differentiation between those with and without psychotic symptoms allows one to hypothesize about potential markers of risk/vulnerability for psychosis versus of disease.

In the current study, we therefore used EEG and a visual adaptation paradigm previously used by our research group to study early visual processing (27, 57), to examine visual processing in a 22q11.2DS group with and without psychotic symptoms, when compared to both an idiopathic schizophrenia cohort and a non-psychiatric control group. Analyses focused on the early visual ERP components evoked by this paradigm during the first ∼200 ms after stimulus presentation. Early visual-evoked responses have been shown to be reduced in schizophrenia (e.g., (27, 33, 58-61)), but see (61-63) for differing evidence. In 22q11.2DS, when no distinction was made between those with and without psychotic symptoms, a combination of reductions and increases was reported in these early time windows (53, 54). This relatively limited ERP evidence for atypical visual processing in 22q11.2DS is strengthened by neuroimaging studies in this population suggesting structural and functional differences in regions associated with visual processing (64, 65). Of note, these regions overlap with areas also affected in schizophrenia (65).

The visual adaptation paradigm utilized here included blocks of different interstimulus intervals (ISIs), thus allowing us to investigate not only basic visual processing, but also sensory adaptation (27). Sensory adaptation is an important property of sensory processing argued to reflect mechanisms by which systems attenuate redundancy (66-69). The amplitude reductions that are typically observed for faster versus slower presentation rates may be due to temporal limitations intrinsic to the mechanisms underlying brain response generation, i.e., faster presentations of stimuli do not allow for full recovery, which results in a decrease of amplitude (70-75). Other explanations have been proposed, such as adaptation as a correlate of priming and/or expectation (76). In schizophrenia, shallower visual adaptation effects have been reported (27, 77, 78), but not consistently (79). In 22q11.2DS, we have previously observed increased auditory adaptation effects in those without psychotic symptoms and decreased auditory adaptation in those with psychotic symptoms (55) when compared to age-matched controls, but visual adaptation has not been investigated in this population.

Given that the presence of psychosis appears to modulate early visual-evoked potentials and that adaptation is diminished in schizophrenia, we hypothesized that there would be differences between those with 22q11.2DS and psychotic symptoms and those with the deletion without psychotic symptoms, reflecting the effect of psychotic symptomatology on neural function. The presence versus absence of such differences would allow us to argue for the usefulness of early visual processing as a marker for psychosis—either of vulnerability or illness. If dysfunctional early visual processing is a marker of risk for schizophrenia, differences should be found between 22q11.2DS (regardless of the presence of psychotic symptoms) and controls, but perhaps not between 22q11.2DS and schizophrenia; if, instead, differences in visual processing are a marker of disease (i.e., psychosis) or illness severity/chronicity, differences should be observed between the two 22q11.2DS groups (and perhaps between each of those groups and the schizophrenia and control groups).

## Materials and Methods

### Participants

Thirty-five individuals with 22q11.2DS (22q; age range: 8-35 years old; 16 with at least one psychotic symptom) and 23 individuals diagnosed with schizophrenia (SZ; age range: 18-63 years old) were recruited. Given the age differences between the two groups, we formed two control groups: one age-matched to the 22q11.2DS sample (CT 22q; N=36, age range: 8-36 years old); the other age-matched to the schizophrenia sample (CT SZ; N=32, age range: 17-62 years old). Individuals with 22q11.2DS were recruited via social media and the Montefiore-Einstein Regional Center for 22q11.2 Deletion Syndrome. All individuals had confirmed genetic diagnosis. Detailed deletion data were available for 20 out of the 35 individuals recruited: 90% presented a large (A-D) deletion. Individuals diagnosed with schizophrenia were recruited through referrals from clinicians in the Department of Psychiatry at Montefiore and Jacobi Health Systems. The recruitment of controls was primarily done by contacting individuals from a laboratory-maintained database and through flyers. Exclusionary criteria for the control groups included developmental and/or educational difficulties or delays, neurological problems, and a severe mental illness diagnosis. Exclusionary criteria for the 22q11.2DS and the schizophrenia groups included current neurological problems. All participants had normal or corrected to normal vision and were asked, at the start of the EEG paradigm, if they could see the fixation cross as well as the color change. All participants signed an informed consent approved by the Institutional Review Board of the Albert Einstein College of Medicine and were monetarily compensated for their time. All aspects of the research conformed to the tenets of the Declaration of Helsinki.

### Experimental Procedure and Stimuli

Testing was carried out over a two-day period and included cognitive testing and EEG recording. IQ measures were computed from the Wechsler Adult Intelligence Scale (WAIS-IV) (80), or the Wechsler Intelligence Scale for Children (WISC-V) (81), depending on age. To assess for the presence of psychotic symptoms, either the Structured Clinical Interview for DSM-5 (SCID-5) (82) or the Structured Clinical Interview for DSM-IV Childhood Diagnoses (Kid-SCID) (83), was performed.

Participants sat in a darkened sound-attenuated electrically shielded booth (Industrial Acoustics Company, Bronx, NY, USA), 65 cm from the screen. They were presented with two blocks of 100% contrast black and white checkerboard annuli (6.5□cm outer diameter and 4.3 cm inner diameter, 1□cm width, 4° × 4°, white luminance of 120□cd□m^−2^, black luminance of 0.2□cd□m^−2^) centered against a gray (luminance = 25 cd m^-2^) background (see Figure 1). Checkerboards were presented for 33 ms, at different ISIs, with a refresh rate of 60 frames/second. To investigate differences in sensory load and in adaptation to repetitive stimulation, five ISIs were used: 145, 245, 495, 995, 2495 ms. A fixation cross was always centrally present, including during checkerboard presentation. Every 10 to 20 seconds, its color changed from blue to yellow for 33 ms and then back to blue again, for an average of 54 color changes. To ensure fixation, participants were instructed to press the right mouse button as soon as th color change was detected. The presentation of checkerboards was temporally unrelated to the fixation task. Between-block intervals were self-paced within a 2500–5000 ms period.

**Figure 1.**
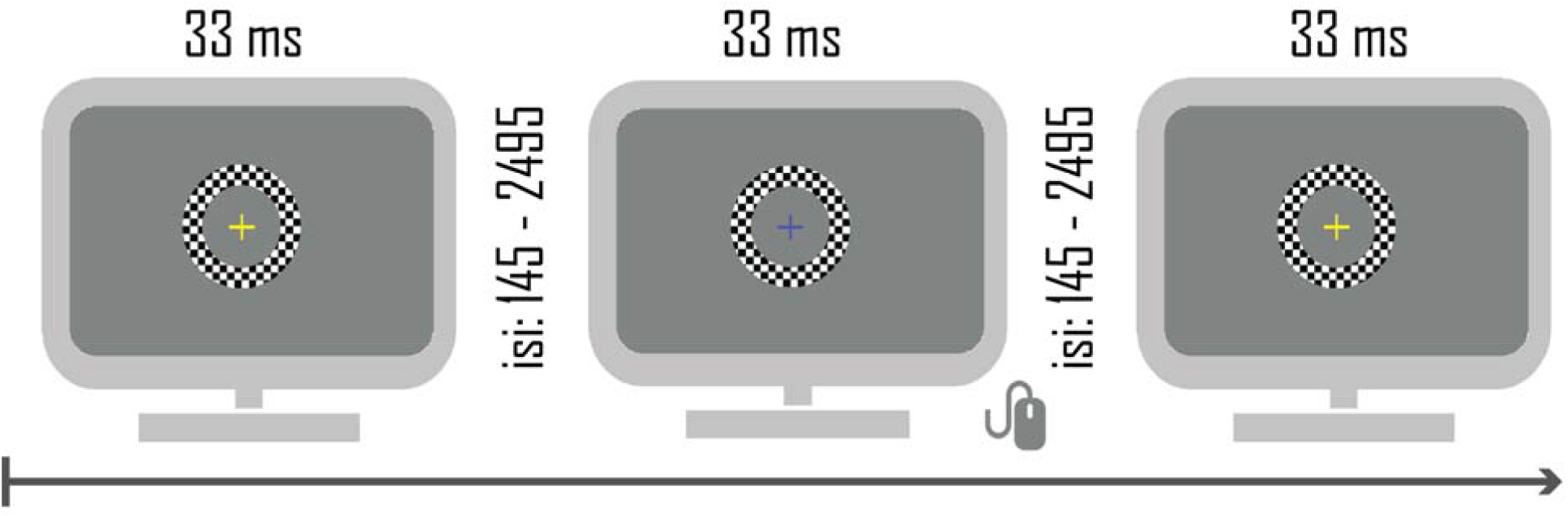
Visual adaptation task. Checkerboards were presented for 33 ms. In five blocked conditions, the ISI was either 145, 245, 495, 995, or 2495 ms.

### Data Acquisition and Analyses

EEG data were acquired continuously at a sampling rate of 512 Hz from 64 scalp locations using electrodes mounted in an elastic cap (Active 2 system; Biosemi™, The Netherlands; 10-20 montage). Preprocessing was carried out using the EEGLAB (version 2019.1) (84) toolbox for MATLAB (version 2019; MathWorks, Natick, MA) (the full pipeline can be accessed at: https://github.com/DouweHorsthuis) (85). Data were downsampled to 256 Hz, re-referenced to the average and filtered using a 1 Hz high pass filter (0.5 Hz transition bandwidth, filter order 1690) and a 45 Hz low pass filter (11 Hz transition bandwidth, filter order 152). Both were zero-phase Hamming windowed sinc finite impulse response (FIR) filters. Bad channels were automatically detected based on kurtosis measures and rejected after visual confirmation. Ocular artifacts were removed by running an Independent Component Analysis (ICA). After ICA, the previously excluded channels were interpolated, using the spherical spline method. Data were segmented into epochs of -50 ms to 400 ms using a baseline of -50 ms to 0 ms. An automatic artifact rejection criterion (moving window peak-to-peak threshold at 120 μV) was applied. Two individuals with 22q11.2DS had a trial exclusion rate higher than 30% and were thus excluded. The number of trials included in the analyses did not differ between groups: CT 22q: 740-990 trials, 22q: 677-986 trials, *p* = .09; CT SZ: 828-989 trials, SZ: 819-992 trials, *p* = .70.

Following Andrade et al. (27), the analysis was focused on central and lateral occipital channels (here, O1, Oz and O2), where signal was maximal in the current data. Windows of interest were between 90 and 140 ms and between 165 and 205 ms, corresponding to the first most prominent peaks generated by this specific paradigm and clearly observed across groups. Trial-by-trial mean amplitude was extracted per time window of interest, channel, and subject. An average of the three channels was used for the statistical analyses. Number of button presses were counted per subject. The experimenter closely monitored the live capture made by the camera inside the booth and, when necessary, reminded the participants to maintain fixation and press the button. Still, as a group, individuals with schizophrenia pressed fewer times than their age-matched peers (CT SZ: M=51.35, SD=3.08, SZ: M=46.34, SD=8.94, *p* = .02). Four individuals with schizophrenia pressed, on average, 30 times, which departed more than one standard deviation from the group average. To investigate the weight of these four individuals in the group average, we compared the amplitude of their brain responses to those of the individuals with schizophrenia who pressed more regularly. As can be seen in Supplementary Figure 1, these two groups did not differ in their visual responses, indicating that those who did not press as often were still fixating. Number of button presses did not differ between individuals with 22q11.2DS and their controls (CT 22q: M=51.64, SD=3.84, 22q: M=50.66, SD=4.65, *p* = .35).

We employed two levels of analyses. First, the 22q11.2DS and schizophrenia groups were compared to their respective age-matched control groups. Mixed-effects models were implemented separately per group (CT 22q *versus* 22q; CT SZ *versus* SZ) to analyze trial-by-trial data, using the *lmer* function in the *lme4* package (86) in R (87). Group and ISI were fixed factors. Participants and trial were added as random factors. Models were fit using the maximum likelihood criterion. *P* values were estimated using *Satterthwaite* approximations. Second, we divided the 22q11.2DS group into two sub-groups: individuals with at least one (positive) psychotic symptom (22q+, N=16) and individuals without psychotic symptoms (22q-, N=17), and compared them. The younger individuals in the 22q-group (N=4, 8-10 years old) were excluded from this analysis, to ensure that these two groups were age-matched. Mixed-effects models were implemented as above. *P-values* from *t*-tests and post-hoc analyses were submitted to Holm-Bonferroni corrections for multiple comparisons (88), using the *p.adjust* of the *stats* package in R (87).

## Results

### Demographics and cognitive function

Table 1 shows a summary of the included participants’ age, biological sex, and IQ. Two-sample independent-means *t* tests were run in R (87) to test for group differences in age and IQ. In cases in which the assumption of the homogeneity of variances was violated, *Welch* corrections were applied to adjust the degrees of freedom. There were more females than males in the CT 22q and 22q groups and more males than females in the CT SZ and SZ groups, but no differences in biological sex between groups. Likewise, age did not differ between the CT 22q and 22q groups and the CT SZ and SZ groups. No differences in sex or age were found between the 22q- and the 22q+ groups. Lower IQs were observed in 22q11.2DS and schizophrenia groups when compared to the control groups. No differences in IQ were observed between 22q- and 22q+.

**Table 1.** Characterization of the controls, 22q11.2DS (overall and per sub-group: 22q- and 22q+), and schizophrenia groups included in the analyses: age, biological sex, and IQ.

|  | age | biological sex | IQ |
| --- | --- | --- | --- |
| **CT 22q** | M=20.00; SD=8.11 | 15 M, 21 F | M=111.67; SD=12.94 |
| **22q** | M=19.48; SD=7.74 | 12 M, 21 F | M=68.81; SD=14.14 |
| t-test | $t=0.27, df=68.98, p=.78$ | - | $t=12.36, df=58.82, p=.01$ |
| Cohen's d | $d=0.06$ | - | $d=3.16$ |
| **22q-** | M=21.62; SD=7.84 | 6 M, 7 F | M=71.91; SD=14.35 |
| **22q+** | M=20.31; SD=6.44 | 4 M, 12 F | M=68.60; SD=13.67 |
| t-test | $t=0.48, df=23.19, p=.63$ | - | $t=0.59, df=21.08, p=.56$ |
| Cohen's d | $d=0.18$ | - | $d=0.24$ |
| **CT SZ** | M=40.34; SD=12.73 | 18 M, 14 F | M=109.27; SD=15.13 |
| **SZ** | M=43.43; SD=11.78 | 16 M, 7 F | M=76.84; SD=8.69 |
| t-test | $t=-0.92, df=49.62, p=.36$ | - | $t=9.52, df=46.67, p=.01$ |
| Cohen's d | $d=0.25$ | - | $d=1.72$ |

Across the four groups, 52.4% reported being white, 20.6% Black or African American, 9.5% as having multiple races, 6.3% Asian, 1.6% American Indian or Native Hawaiian, and <1% Hawaiian or Pacific Islander. 7.9% reported unknown racial background or declined to answer. 19.1% of the individuals tested were Hispanic or Latino and 73.8% non-Hispanic or non-Latino. 6.3% reported unknown ethnic background or declined to answer.

17.1% of those with 22q11.2DS were diagnosed with a mood disorder, 17.1% with an anxiety disorder, 2.8% with a conduct disorder, 5.7% with an attention disorder, and 8.6% with schizophrenia. In the schizophrenia group, 8.7% met criteria for a mood disorder and 8.7% for an anxiety disorder. 22.8% of those with 22q11.2DS were taking antidepressants, 28.6% anticonvulsants, 11.4% antipsychotics, 5.7% antimanics, and 11.4% stimulants. From those individuals diagnosed with schizophrenia, 17.4% were taking antidepressants, 21.8% anticholinergics, and 21.8% anticonvulsants. All of those with schizophrenia were taking antipsychotics. One individual with schizophrenia was taking testosterone. In the control groups, 2.7% reported a mood disorder and 5.6% an anxiety disorder. 5.6% were taking antidepressants.

### Event-related potentials and Adaptation

#### 22q11.2DS and Controls

Figure 2 depicts averaged ERPs from occipital channels O1, Oz and O2 and topographies for CT 22q and 22q groups.

**Figure 2.**
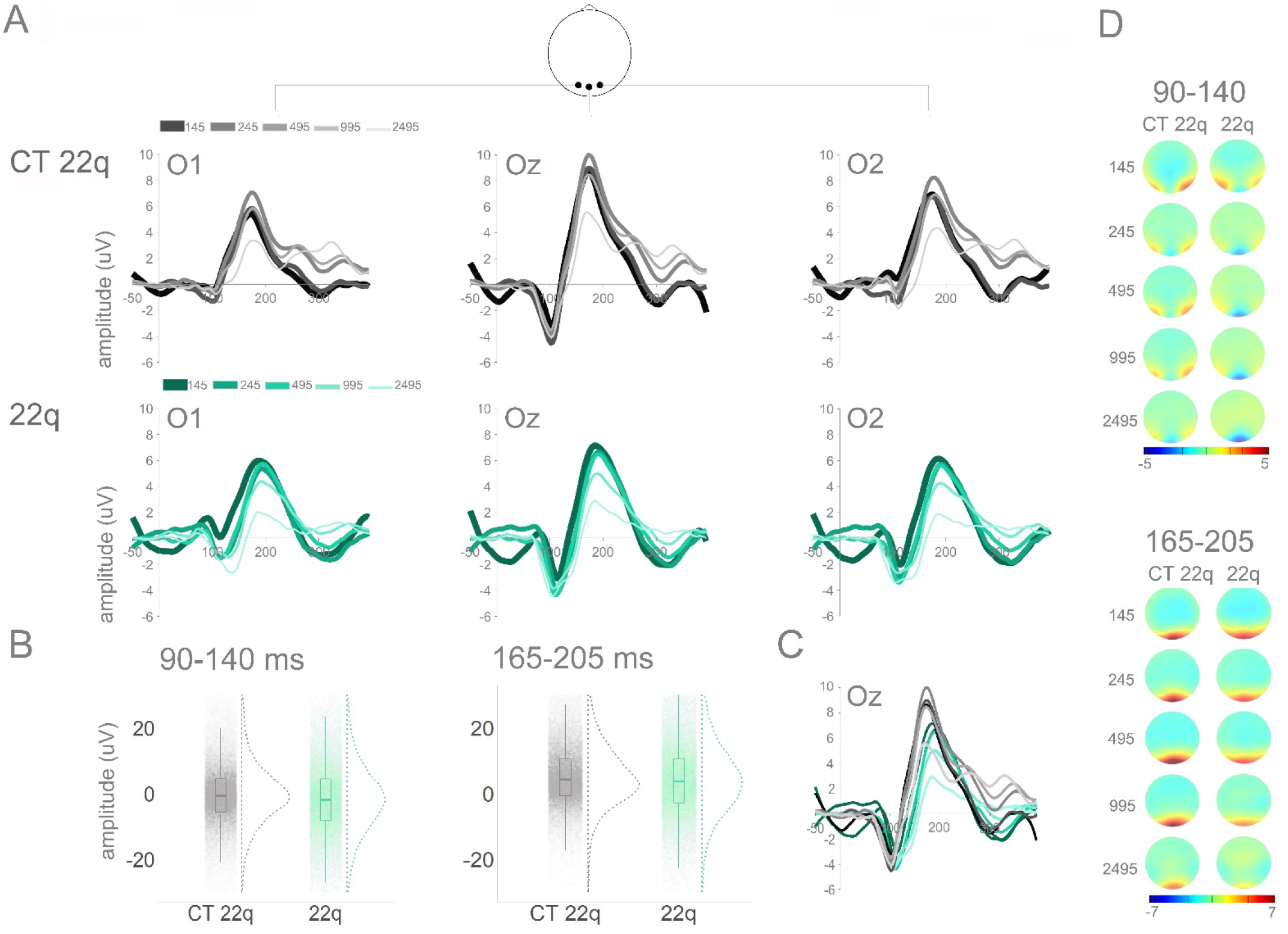
Panel A: Averaged ERPs per group (CT 22q and 22q) and ISI at O1, Oz, and O2. Panel B: Plots showing distribution of amplitudes (trial-by-trial data; average of O1, Oz, and O2) for the time windows of interest (90-140 ms and 165-205 ms). Panel C: Topographies for windows of interest per group and ISI.

In the 90-140 ms time window, though no significant effect of group was found (ß = -1.03, SE = 0.98, *p* = .29), there was a significant interaction between group and ISI: When compared to their control group, individuals with 22q11.2DS showed more negative amplitudes for the 245 (ß = -0.65, SE = 0.25, *p* = .01), the 495 (ß = -1.54, SE = 0.25, *p* = .001), the 995 (ß = -1.17, SE = 0.25, *p* = .001), and the 2495 (ß = -0.95, SE = 0.25, *p* = .001) ISIs when compared to the shortest ISI (145), suggesting larger adaptation in those with 22q11.2DS (see Figure 3A). Post-hoc tests revealed that, when compared to their controls, individuals with 22q11.2DS presented particularly increased amplitudes in the 495 (ß = -2.51, SE = 0.99, *p* = .01) and in 995 (ß = -2.23, SE = 1.12, *p* = .04) ISIs (Figure 2A). As can be appreciated in Figure 2, this response appears to be more diffuse in 22q11.2DS when compared to their controls, who showed a more focal (central) response. There was also an effect of ISI. Across groups, 145 ms resulted in decreased amplitudes when compared to the remainder ISIs: 245 (ß = -1.00, SE = 0.17, *p* = .001); 495 (ß = -0.34, SE = 0.17, *p* = .04); 995 (ß = -0.76, SE = 0.17, *p* = .001); 2495 (ß = -1.78, SE = 0.17, *p* = .001) (Figures 2 and 3A).

**Figure 3.**
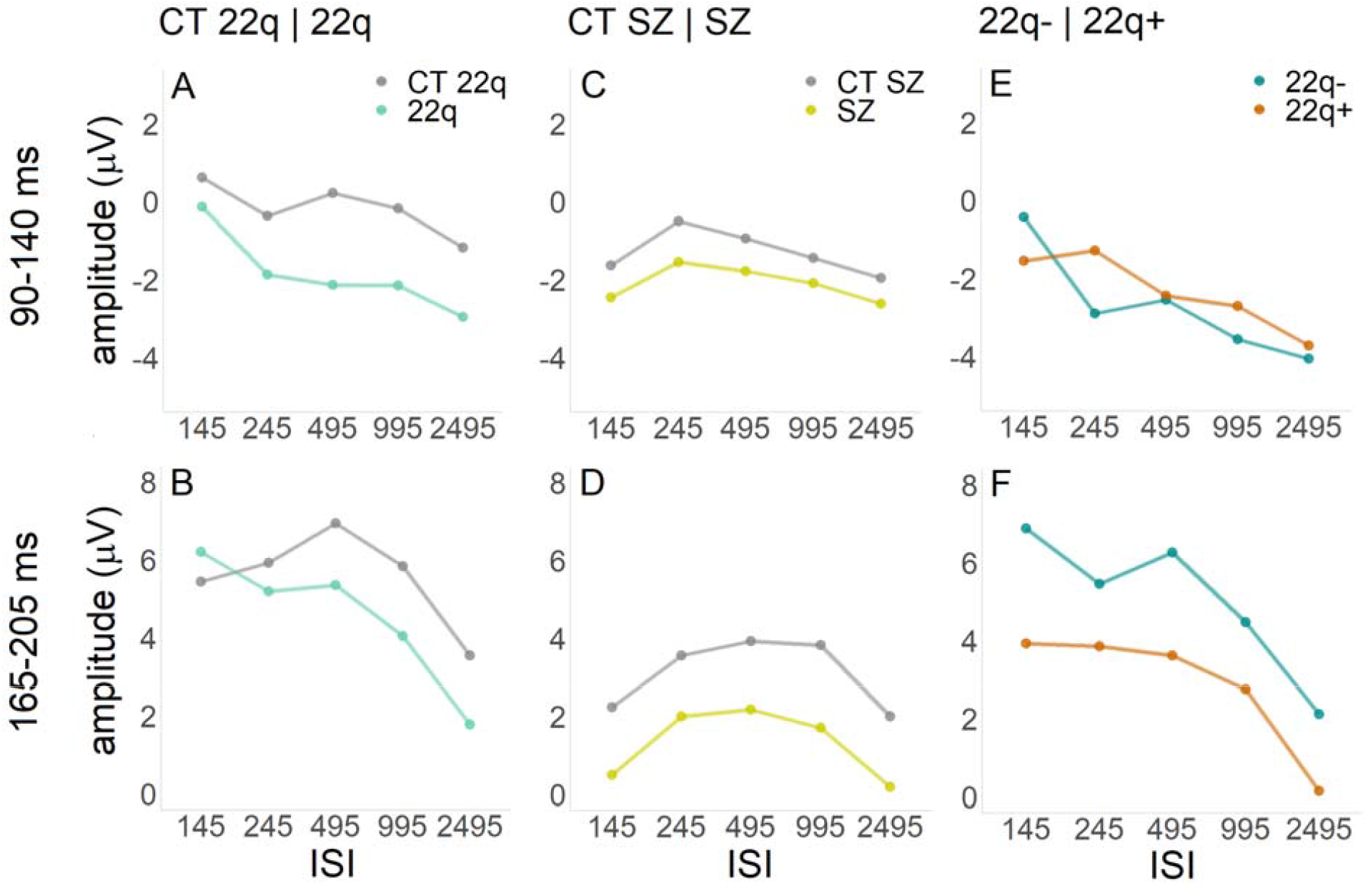
Curves representing visual adaptation effects between groups.

As one can observe in Figure 2, amplitudes in the 165-205 ms time window appear to be slightly decreased in those with 22q11.2DS when compared to their controls, particularly at the longest ISIs. These apparent differences were not, however, statistically significant (ß = 0.84, SE = 1.04, *p* = .42). Once again, the interaction between group and ISI was significant. When compared to the CT 22q group, the 22q group showed more negative amplitudes for the 245 (ß = -1.55, SE = 0.26, *p* = .001), the 495 (ß = -2.34, SE = 0.26, *p* = .001), the 995 (ß = -2.56, SE = 0.26, *p* = .001), and the 2495 (ß = -2.57, SE = 0.26, *p* = .001) ISIs when compared to the shortest ISI (145), again suggesting more sensitive adaptation in those with 22q11.2DS (see Figure 3B). There was also an effect of ISI. The 245 (ß = 0.50, SE = 0.17, *p* = .01), 495 (ß = 1.54, SE = 0.17, *p* = .001), and 995 (ß = 0.47, SE = 0.17, *p* = .01) ISIs resulted in more positive amplitudes when compared to the 145 ISI. In contrast, the 2495 ISI was more negative than the 145 ISI (ß = -1.86, SE = 0.17, *p* = .001). As can be appreciated in Figure 3B, this ISI effect is primarily explained by the control group.

### Schizophrenia and Controls

Figure 4 depicts averaged ERPs from occipital channels O1, Oz and O2 and topographies for CT SZ and SZ groups. Overall, individuals with schizophrenia, when compared to their control peers, present similar amplitudes in the earlier window of interest (90-140 ms), but reduced amplitudes in the later window of interest (165-205 ms). Additionally, as in 22q11.2DS, those with schizophrenia appear to show a less focal response when compared to their age-matched control group.

**Figure 4.**
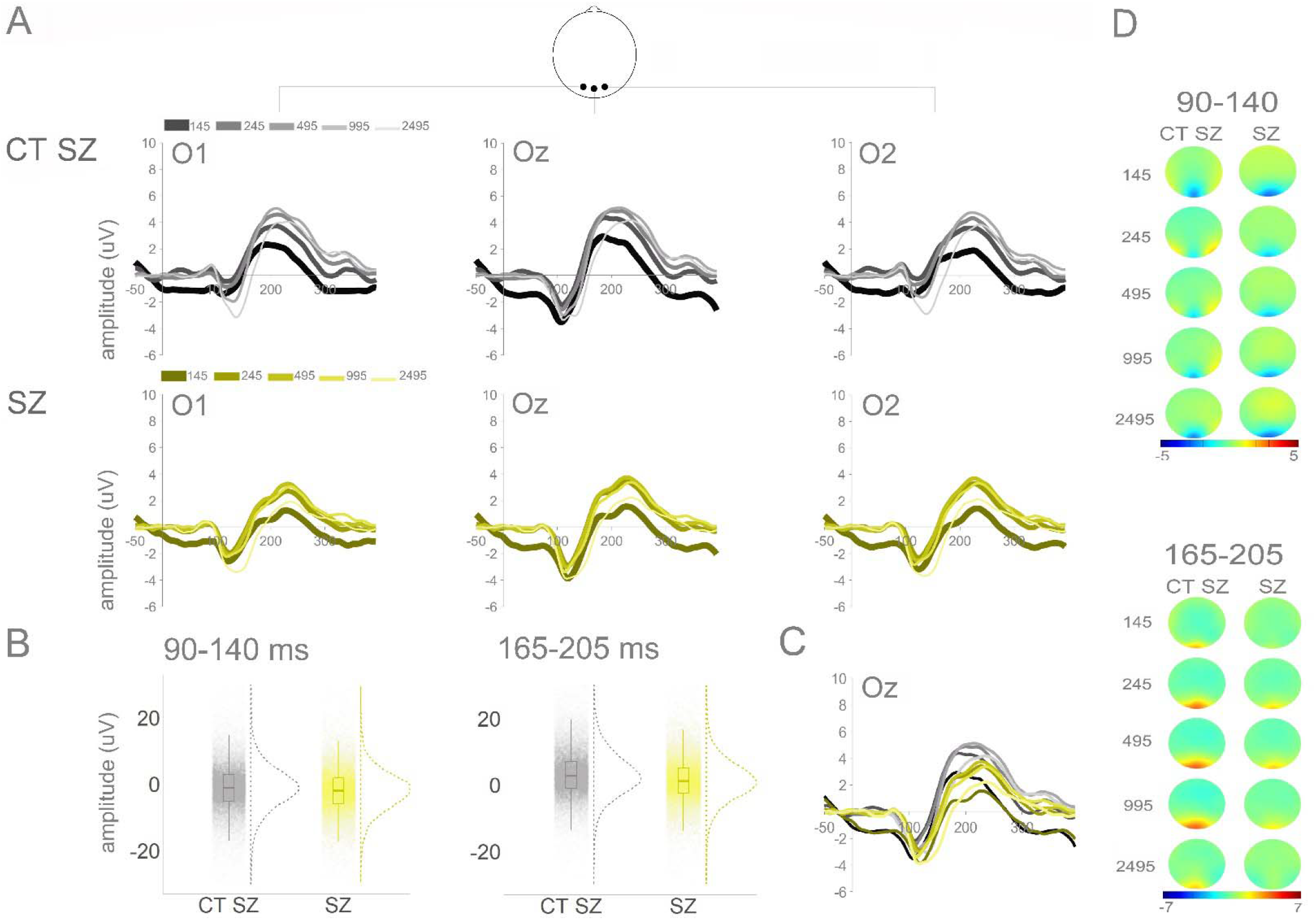
Panel A: Averaged ERPs per group (CT SZ and SZ) and ISI at O1, Oz, and O2. Panel B: Plots showing distribution of amplitudes (trial-by-trial data; average of O1, Oz, and O2) for the time windows of interest (90-140 ms and 165-205 ms). Panel C: Topographies for windows of interest per group and ISI.

In the 90-140 ms time window, no significant effect of group (ß = -0.88, SE = 0.65, *p* = .19) or of the interaction between group and ISI were found. To more thoroughly explore the spatio-temporal dynamics of these responses, a post-hoc statistical cluster plot was computed (Figure S2 in the Supplementary Material). This plot suggests that there may be slight differences between individuals with schizophrenia and their controls in occipital channels staring at around 100ms. There was an effect of ISI in the 90-140 ms time window: The 245 (ß = 1.12, SE = 0.11, *p* = .001) and the 495 (ß = 0.69, SE = 0.11, *p* = .001) ISIs were more positive when compared to the 145 ISI; the 2495 ISI was more negative when compared to the 145 ISI (ß = -0.33, SE = 0.11, *p* = .01) (see Figure 3C).

In the 165-205 ms time window, there was an effect of group, with individuals with schizophrenia showing reduced amplitudes compared to controls (ß = -1.66, SE = 0.69, *p* = .02). There was also a significant interaction between group and ISI, with the SZ group presenting a smaller difference between the 145 and the 995 ISIs, when compared to controls (ß = -0.51, SE = 0.18, *p* = .01) (see Figure 3D). Lastly, and as can be observed in Figure 3D, there was an effect of ISI. The 245 (ß = 1.30, SE = 0.12, *p* = .001), 495 (ß = 1.71, SE = 0.12, *p* = .001), and 995 (ß = 1.65, SE = 0.12, *p* = .001) ISIs resulted in more positive amplitudes when compared to the 145 ISI.

### 22q with and without psychotic symptoms

Figure 5 depicts averaged ERPs in occipital channels O1, Oz and O2 and topographies for 22q- and 22q+ groups. Overall, individuals with 22q11.2DS and at least one psychotic symptom (22q+), when compared to those with 22q11.2DS but no psychotic symptoms, present reduced amplitudes.

**Figure 5.**
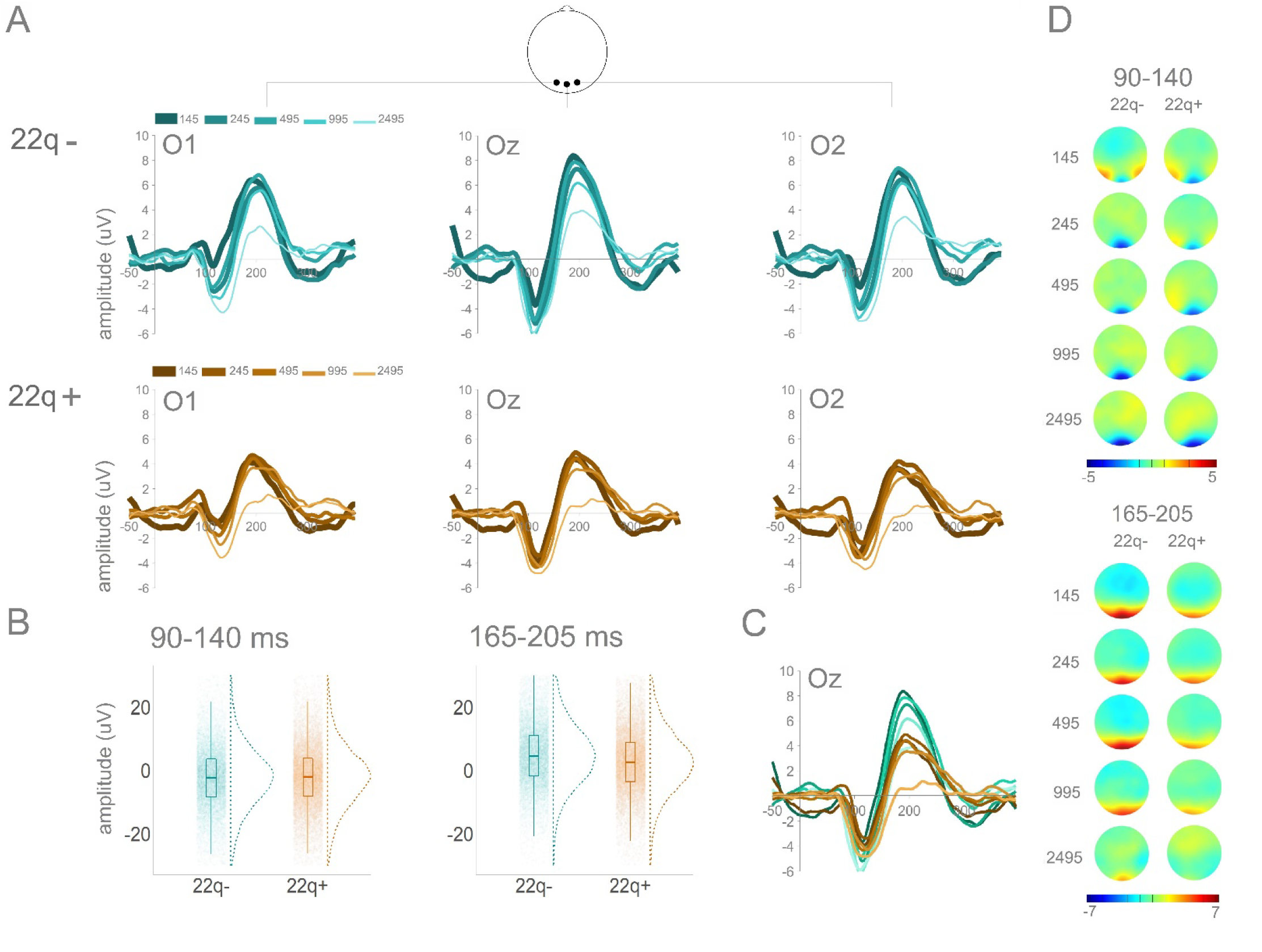
Panel A: Averaged ERPs per group (22q- and 22q+) and ISI at O1, Oz, and O2. Panel B: Plots showing distribution of amplitudes (trial-by-trial data; average of O1, Oz, and O2) for the time windows of interest (90-140 ms and 165-205 ms). Panel C: Topographies for windows of interest per group and ISI.

In the 90-140 ms time window, though no significant effect of group was found (ß = -1.12, SE = 1.32, *p* = .40), there was a significant interaction between group and ISI: When compared to the 22q-group, the 22q+ group showed less negative amplitudes for the 245 (ß = 2.61, SE = 0.40, *p* = .001), the 495 (ß = 1.17, SE = 0.40, *p* = .004), the 995 (ß = 2.05, SE = 0.40, *p* = .001), and the 2495 (ß = 1.52, SE = 0.40, *p* = .001) ISIs compared to the shortest ISI (145) (see Figure 3E). There was also an effect of ISI. Across groups, 145 ms resulted in less negative amplitudes when compared to the other ISIs: 245 (ß = - 2.31, SE = 0.30, *p* = .001); 495 (ß = -1.96, SE = 0.30, *p* = .001); 995 (ß = -3.06, SE = 0.30, *p* = .001); 2495 (ß = -3.54, SE = 0.30, *p* = .001) (Figure 3E).

In the 165-205 ms time window, there was an effect of group: 22q+ showed decreased amplitudes (ß = -3.07, SE = 1.31, *p* = .03) when compared to 22q-. As one can appreciate in Figure 3F, there was also a significant interaction between group and ISI, with the 22q+ group presenting reduced differences between the 145 and the 245 (ß = 1.38, SE = 0.42, *p* = .001), the 995 (ß = 0.93, SE = 0.43, *p* = .03), and the 2495 (ß = 0.84, SE = 0.43, *p* = .049) ISIs, when compared to the 22q-group. Lastly, there was an effect of ISI. The 245 (ß = -1.42, SE = 0.32, *p* = .001), 995 (ß = -2.13, SE = 0.32, *p* = .001), and 2495 (ß = -4.67, SE = 0.32, *p* = .001) ISIs resulted in more negative amplitudes when compared to the 145 ISI (Figure 3F).

In summary, and as can be appreciate in Figure 6A, there appears to be a continuum of amplitudes between CT 22q and schizophrenia, with those with 22q11.2DS without and with symptoms lying in between those two groups. Figure 6B summarizes the adaptation effects for the time windows of interest: 90-140 ms and 165-205 ms. While in the former, differential patterns are less clear, in the latter, and when comparing the amplitudes evoked by either the two most extreme ISIs (145 vs. 2495), or by the smallest versus biggest amplitude response across the five ISI conditions, both 22q groups present increased adaptation when compared to age-matched controls and individuals with schizophrenia.

**Figure 6.**
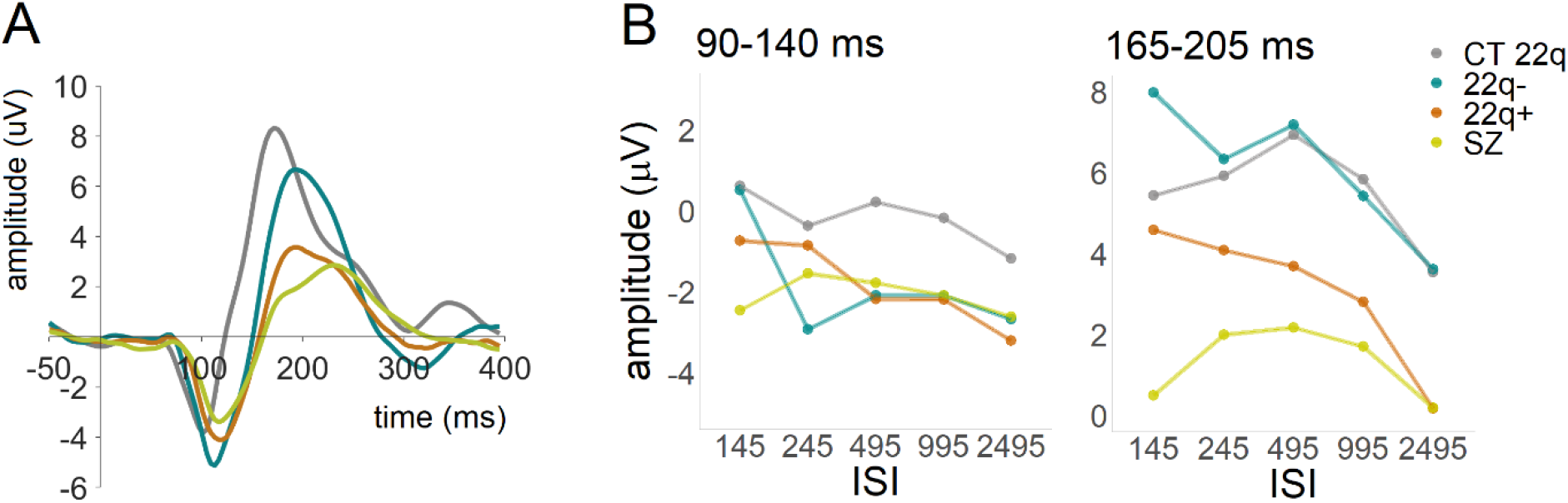
Panel A: Averaged ERPs per group (CT 22q, 22q-, 22q+, and SZ) at Oz, showing the average of all ISIs. Panel B: Curves representing visual adaptation effects between groups (CT 22q, 22q-, 22q+, and SZ) in the time windows of interest (90-140 ms and 165-205 ms).

## Discussion

Utilizing EEG and a visual adaptation task, we investigated early visual processing and adaptation in a sample of individuals with 22q11.2DS with and without psychotic symptoms and compared their brain responses to control and schizophrenia groups. Given the high risk for schizophrenia in 22q11.2DS, we expected to see similarities in the pattern of brain responses and sensory adaptation between 22q11.2DS, particularly those with psychotic symptoms, and schizophrenia. While all groups exhibited robust visual-evoked potentials of generally similar morphology and showed similar patterns of adaptation, significant differences were present between clinical and control groups, and between th 22q11.2DS groups. Similar to our findings in the auditory domain (55), ERPs at ∼100 ms were increased in 22q-compared to the 22q+ and control groups. This emphasizes the thesis that there are significant differences in neural function in 22q11.2DS and stresses the importance of distinguishing between thos with 22q11.2DS and psychotic symptoms and those without. Subsequent visual processing also differed as a function of group, with the schizophrenia and 22q+ groups showing decreased amplitudes in the 165-250 ms time window when compared to the control and 22q-groups, respectively. These significant group differences suggest basic mechanistic and disease process effects on neural processing that are evident in early visual processing, with possible implications for phenotype.

In considering our findings in greater detail and within the context of the extant literature, due to somewhat atypical EEG responses evoked by the stimuli used in our visual adaptation paradigm (more centrally focused than typical and of reversed directionality) (27, 57), here, we focus on the timeframes of the major responses, rather than on the classic P1/N1 nomenclature.

A previous study on visual processing in 22q11.2DS reported early (∼100 ms) visual processing reductions (53). In that study, however, no distinction was made between those with and without psychotic symptoms, a distinction that we have shown to be highly relevant (55, 56). And indeed, here, the increased amplitudes between 90 and 140 ms in 22q11.2DS appear to be mainly driven by those without psychotic symptoms (see Figures 5 and 6A). Notably, and in accord with what we report here, though not distinguishing between individuals with and without psychotic symptoms, Biria et al. argued that individuals with 22q11.2DS who have a lower risk for schizophrenia might present an increased and compensatory visual response when compared to controls (53). Interestingly, similar increases were observed in the auditory modality in three human studies (55, 89, 90) and in a mouse model of the deletion (91). Thus, though our results are not entirely consistent with Biria et al., they do align with what has been observed in other sensory modalities. This cross-modality increase in sensory brain responses in 22q11.2DS (particularly in the absence of psychotic symptoms) might thus constitute a marker of the deletion and hold potential to better understand neurogenetic mechanisms.

That no clear differences were found in this earlier time window (90-140 ms) between individuals with schizophrenia and their control peers could be explained by relatively maintained early stages of visual processing in this population. However, differences in early visual-evoked potentials in schizophrenia at ∼100 ms have been consistently reported in the literature (4, 28, 30, 37, 51, 63, 92). Of note, here, though such differences were not captured by our more conservative primary analysis, Figure S2 suggests the presence of slight differences between the two groups starting at around 100ms. That, and the fact that, in our previous work, we have shown significant differences between those with and without schizophrenia utilizing this same paradigm (27), suggests that the visual adaptation paradigm used here may lack sensitivity. By generating small effect sizes, analyses are underpowered, and findings are easily missed. Nevertheless, considering that no differences were found between those with schizophrenia or those with 22q11.2DS and psychotic symptoms and those without, amplitude in this time window, as evoked by this specific paradigm, does not appear to be a meaningful marker for psychosis. It is additionally interesting to note that responses in the 90-140ms window in 22q11.2DS and schizophrenia seem to be less focal when compared to the control groups: Whereas in controls, the signal is maximal centrally, in 22q11.2DS and schizophrenia, it appears to be equally large on the central, left, and right occipital channels. Though differences in topography were not statistically tested here, one possibility which warrants investigation is that there is less specificity of receptive fields in the visual cortex in these clinical populations. Alternatively, there may be underlying anatomical differences between these groups which explain the less focal activity. Additional work is needed to understand the implications of a more diffuse response during early cortical visual processing.

In the 165-205 ms time window, the 22q+ group differed from the 22q-, and those with schizophrenia differed from their controls. Both groups (22q+ and schizophrenia) presented reduced amplitudes. In agreement with these findings, reduced response amplitudes in this time window have been previously shown in 22q11.2DS (53). In schizophrenia, the reported findings are discrepant: While some studies argue for no differences in visual-evoked responses between individuals with and without schizophrenia (37, 62, 93, 94), others have described reduced amplitudes in schizophrenia (38, 95, 96). Of note, these reductions appear to be modulated by the type of task used or the process measured. While studies reporting no differences have focused on illusory contour processing and contour integration (37, 62, 93, 94), those arguing for the presence of reduced amplitudes in schizophrenia in this time window utilized global versus local (95), go/no-go (38), and backward masking (96) tasks. Reduced amplitudes in schizophrenia may therefore be expected in this time window under certain conditions. It is nonetheless interesting that, here, amplitude reductions were likewise observed in individuals with psychotic symptoms (even in the absence of a diagnosis of schizophrenia), but not in those at-risk but with no psychotic symptoms. Reductions in this time window might thus reflect disease, rather than risk/vulnerability for psychosis. Importantly, early visual processing deficits are also seen in bipolar disorder (97), and thus may be ubiquitous to psychotic disorders more generally. Of note, given that the mere presence of psychotic symptoms led to such reductions but that those were exacerbated in the chronic schizophrenia group, brain activity in this time window seems likely to be modulated not just by presence of disease, but also by chronicity or severity. Figure 6 suggests that not only amplitude, but also latency might differ between groups, particularly between 22q- and controls. Taking into consideration the increased early (90-140 ms) amplitudes (also seen in Figure 6) for the 22q-group, this potential difference in latency could be a function of the earlier increased amplitude. Nonetheless, differences in latency should be further explored in future research.

Increased/preserved early (∼100 ms) and decreased later (∼200 ms) responses have been associated with alterations in cortical glutamate N-methyl-D-aspartate (NMDA) receptors (89, 98-100): NMDA-related dysfunctional mechanisms could alter the modulation of sensory information reflected by early components and the efficiency of early attentional processes indexed by later components (100). Increased sensitivity to NMDA receptor antagonism has been described in a mouse model of 22q11.2DS (91) and elevated NMDA-receptor antibodies were found in a 19-year-old individual with the deletion and a history of cognitive decline and psychotic symptomatology (101). Furthermore, NMDA receptor antagonists have been shown to replicate many of the clinical features of schizophrenia (for a review, see (102)). Additionally, basic visual sensory processing differences have already been associated with two NMDA-related genes, *DTNBP1* and *NOS1*, both implicated in schizophrenia risk (103, 104), and haploinsufficiency of PRODH in 22q11.2DS has been proposed as a modifier gene for schizophrenia: By modulating cortical dopaminergic transmission and glutamatergic signaling during early brain development, PRODH could influence vulnerability for schizophrenia (105). A more thorough, longitudinal investigation of these associations and the roles of these genes in the conversion to psychosis in 22q11.2DS, has potential to advance our knowledge about the contribution of specific neural and genetic processes (and of their interactions) to the onset of schizophrenia. This may additionally play a vital role in drug development.

With regard to adaptation, in both time windows, differences were present between 22q11.2DS and controls and between 22q- and 22q+. 22q11.2DS (regardless of the presence of psychotic symptoms) and 22q-groups presented increased adaptation effects. Consistent with this finding, in a previous study investigating basic auditory processing and sensory memory in 22q11.2DS, we showed increased auditory adaptation effects in individuals with the deletion (55). Larger adaptation effects have been argued to reflect better encoding efficiency in the visual modality and to relate to enhanced visual short-term memory and attentional processes (106-108), which would suggest more efficient encoding in 22q11.2DS when compared to controls (and in 22q-when compared to 22q+). Without objective measures of such efficiency, this study does not allow one to draw conclusions in that regard. Worth mentioning, however, is that in a previous study in which working memory was assessed in an identical sample, individuals with 22q11.2DS performed significantly worse in a verbal working memory task and no differences were found between those with and without psychotic symptoms (55). Moreover, difficulties have been noted in visuospatial memory, attention, and other executive-type functions in this population (109-113). Hence, it seems exceedingly unlikely that, here, the increased adaptation effects observed in 22q11.2DS relate to better encoding and memory. The nature of the neural mechanisms underlying visual adaptation and how adaptation propagates through the visual stream are not fully understood and thus little is also known about the potential causes of the enlarged adaptation seen in in 22q11.DS, particularly in those without psychotic symptoms. One study suggests that a downregulation of the GABA-ergic system may lead to auditory adaptation impairments (114). Pharmacological studies targeting neurotransmitter systems implicated in visual adaptation may contribute to a better understanding of this specific type of sensory adaptation. Still, and as for the enlarged amplitude in the 90-140 ms time window, increased adaptation across modalities in 22q11.2DS may be a relevant marker of the deletion and reflect relevant and specific neurogenetic processes with potential clinical implications.

The reduction of adaptation in 22q+ is, likewise, of interest and recapitulates our prediction of blunted adaptation in psychosis. One proposed function of adaptation is that it improves discriminability of novel stimuli (115). Given that one explanation for psychotic symptoms is the inappropriate attribution of salience to irrelevant stimuli, this hypothesis is particularly relevant for schizophrenia (116) and for those with 22q11.2DS and psychotic symptoms, who, in the current study, presented reduced adaptation. Importantly, Silverstein et al. argued that blunted adaptation may underpin differences in the salience ascribed to stimuli (117). And, considering that adaptation, at the very least, ought to allow the visual system to leverage input regularities to heighten visual processing, reduced visual adaptation may contribute to the visual perception differences characteristic in psychosis (118). Future research should attempt to clarify the relationship between adaptation and the presence of psychotic symptoms in specific sensory modalities. Additionally, a better characterization of the behavioral and functional impacts of differences in adaptation is necessary to understand its true value for this clinical population.

That no clear adaptation was observed in schizophrenia could have been explained by psychosis-related pathological processes and would not be wholly unexpected in a chronic schizophrenia sample: Reduced visual adaption effects have been previously described in this population (79). However, the lack of adaption appears to be likewise present in the control group for schizophrenia (see Figure 3). A study investigating the effects of aging in visuomotor adaptation indicated that, when compared to a younger sample, older individuals did not adapt to the same extent (119). Our samples were not, however, as old as the elderly sample tested in that study and a reduced shorter ISI appears to be present in younger controls too (Figure 6), and thus, the effects of aging are an unlikely explanation for our findings. Large samples would, nevertheless, have allowed for further analyses on the impact of age in this result. Alternatively, and considering that this lack of adaptation effects in both controls and individuals with schizophrenia might be in part due to a particularly reduced short (145ms) ISI when compared to all others (Figure 3), we argue that this finding might relate to methodological aspects, rather than to pathological ones. Indeed, Figures 3 and 6 appear to indicate that there was some level of adaption occurring between the longer ISIs for both groups. The problematic baseline for the 145ms ISI, caused by the rate of stimulation, appears to be somehow more prominent in these two groups, and could partially explain the absence of adaptive differences between the shortest and the longer ISIs—at least in the earlier time window (and perhaps in interaction with the effects of age). Importantly, a previous study conducted in our lab investigating visual adaptation in schizophrenia—in which shallower adaptation was found for that clinical population—utilized slightly longer ISIs (i.e., 200ms instead of 145ms), which could explain the differences in the findings reported in the two studies. It is important to stress that, regardless, the differences between individuals with schizophrenia and their control peers are, here, merely in amplitude, not in adaptation. Hence, adaptation effects, though conceivably valuable measures of neuronal plasticity with potential to shed light on psychosis clinical phenomenology, do not seem to distinguish between those with chronic schizophrenia and their controls in the current study. One could note that adaptation may be reduced in schizophrenia when compared to 22q11.2DS (Figure 6), but age differences complicate such comparison.

Some of the limitations of the current study have been made clear throughout the Discussion. Furthermore, we have alluded to a few remaining questions for potential future research. The following additional limitations of this study should be considered. The impacts of medication and co-morbidities were not taken into consideration, given the relatively small samples and the heterogeneity in those variables. With regard to medication, however, visual processing deficits have been found in both medicated and unmedicated individuals (22, 120-122). Age-matched 22q11.2DS and schizophrenia samples would have provided better opportunities to characterize similarities and differences between these populations. Future work should utilize different measures of psychotic symptomatology. Instruments providing symptom severity and the differentiation between negative and positive symptoms and between different sensory modalities would have been more informative. Lastly, attention appears to modulate auditory adaptation (123, 124). Future research should additionally address the effects of attention in visual adaptation (123, 124), given that attentional difficulties are characteristic of both 22q11.2DS and schizophrenia.

Taken together, the present data suggest that while amplitude and adaptation in an earlier time window of visual processing (90-140 ms) may reflect neurogenetic mechanisms associated with a deletion in chromosome 22, amplitude in a later window (165-205 ms) may be a marker of the presence of psychosis and/or of its chronicity/severity. It is nonetheless important to consider that the schizophrenia phenotype associated with 22q11.2DS may be a unique one, and not necessarily related to the idiopathic form of the condition, which is diagnosed by symptom clusters rather than neurobiology. These findings, particularly the cross-modality increase in brain responses and adaptation seen in 22q11.2DS—when also considering Francisco et al (55), may hold important clinical implications for potential treatment development.

## Data Availability

All data produced in the present study are available upon reasonable request to the authors

## Acknowledgements

We wish to thank Dr. Juliana Bates, who performed the clinical assessments, and Alaina Berruti for her help with data collection. Additionally, we thank Chloe Ifrah and Ilana Deyneko for their assistance at different stages of the project. We would also like to acknowledge the role of the Jacobi Medical Center, the Montefiore-Einstein Regional Center for 22q11.2 Deletion Syndrome, and the OnTrackNY program at Montefiore in recruitment, and the Rose F. Kennedy Intellectual and Developmental Disability Research Center for all its support. We extend our most sincere gratitude to the participants and their families for their interest, their involvement, and their time.

This work was supported in part by the Eunice Kennedy Shriver National Institute of Child Health and Human Development (NICHD), under award number U54 HD090260. Work on rare diseases at the University of Rochester (UR) collaboration site is supported in part through the UR Intellectual and Developmental Disabilities Research Center (UR-IDDRC), which is funded by a center grant from the Eunice Kennedy Shriver National Institute of Child Health and Human Development (NICHD P50 HD103536 to JJF).

## Author contributions

JJF, SM, and AAF conceived the study and designed the original experiment. AAF and DJH collected and analyzed the data. AAF wrote the first draft of the manuscript. SM, JJF, and DJH provided editorial input on the subsequent drafts. All authors read and approved the final manuscript.

## Data accessibility statement

Data can be accessed upon request. Analysis pipeline can be accessed at github.com/DouweHorsthuis.

## Supplementary material

**Figure S1.**
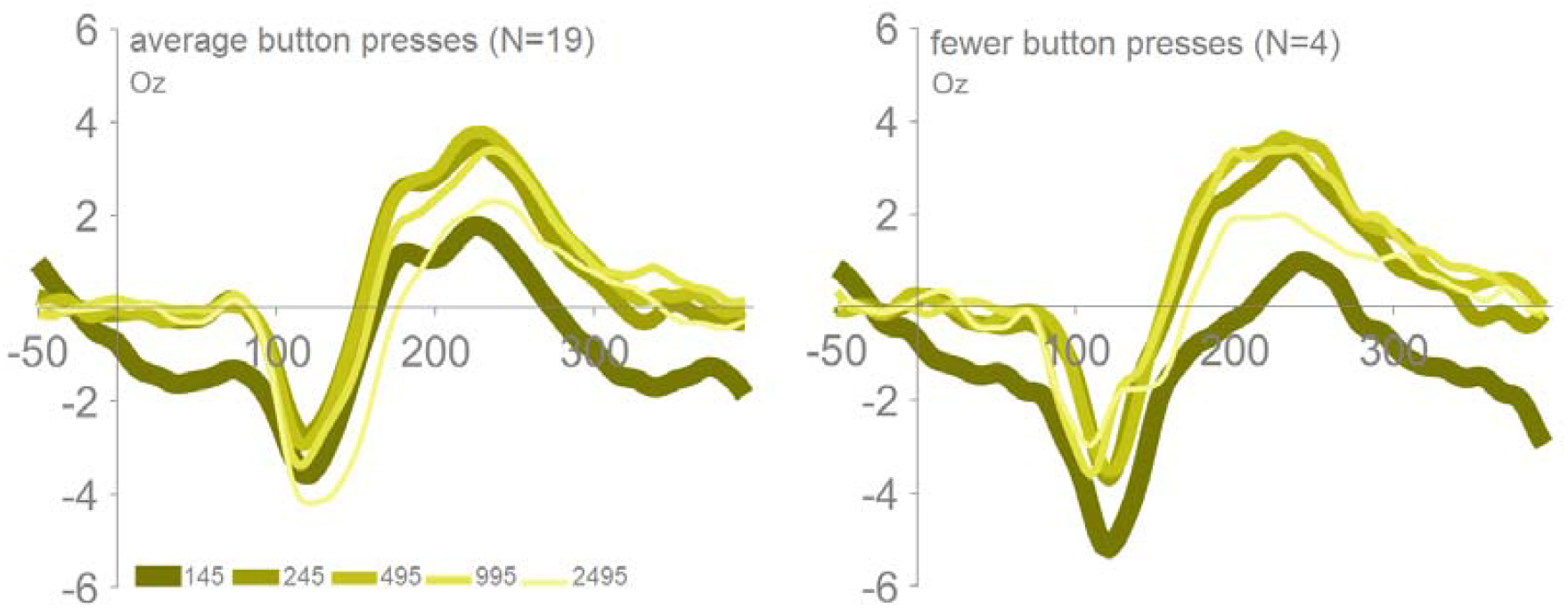
Averaged ERPs per ISI at Oz, showing VEPs for individuals with schizophrenia with average number of button presses versus those with schizophrenia and significantly fewer button presses.

**Figure S2.**
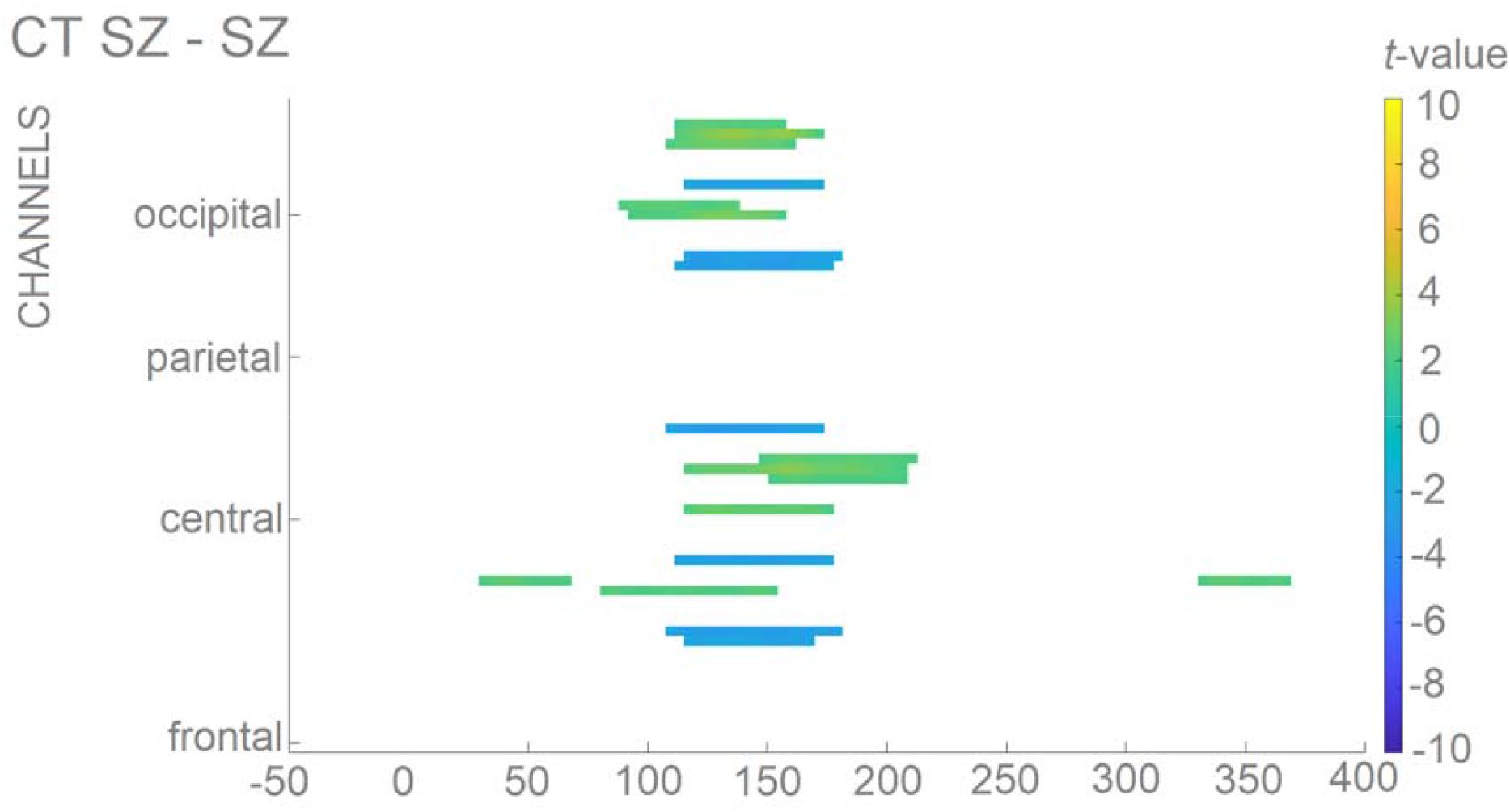
Statistical cluster plot. Color values indicate the p-values that result from point-wise *t*-tests evaluating the responses across time (*x*-axis) and electrode positions (*y*-axis). General electrode positions are arranged from frontal to occipital regions (bottom to top) and the scalp has been divided into four general scalp regions. Within each general region, electrode laterality is arranged from left to right Only *p*-values <0.05 are color-coded.

## References

1. Baker KD, Skuse DH (2005): Adolescents and young adults with 22qll deletion syndrome: psychopathology in an at-risk group. The British Journal of Psychiatry. 186:115–120.

2. Monks S, Niarchou M, Davies AR, Walters JT, Williams N, Owen MJ, et al. (2014): Further evidence for high rates of schizophrenia in 22q11. 2 deletion syndrome. Schizophrenia Research. 153:231–236.

3. Bassett AS, Chow EW (1999): 22q11 deletion syndrome: A genetic subtype of schizophrenia. Biological Psychiatry. 46:882–891.

4. Butler PD, Javitt DC (2005): Early-stage visual processing deficits in schizophrenia. Current Opinion in Psychiatry. 18:151–157.

5. Yoon JH, Maddock RJ, Rokem A, Silver MA, Minzenberg MJ, Ragland JD, et al. (2010): GABA concentration is reduced in visual cortex in schizophrenia and correlates with orientation-specific surround suppression. Journal of Neuroscience. 30:3777–3781.

6. Brenner CA, Wilt MA, Lysaker PH, Koyfman A, O’Donnell BF (2003): Psychometrically matched visual-processing tasks in schizophrenia spectrum disorders. Journal of Abnormal Psychology. 112:28–37.

7. Chen Y, Nakayama K, Levy D, Matthysse S, Holzman P (2003): Processing of global, but not local, motion direction is deficient in schizophrenia. Schizophrenia Research. 61:215–227.

8. Chen Y, Palafox GP, Nakayama K, Levy DL, Matthysse S, Holzman PS (1999): Motion perception in schizophrenia. Archives of General Psychiatry. 56:149–154.

9. Stuve TA, Friedman L, Jesberger JA, Gilmore GC, Strauss ME, Meltzer HY (1997): The relationship between smooth pursuit performance, motion perception and sustained visual attention in patients with schizophrenia and normal controls. Psychological Medicine. 27:143–152.

10. Tadin D, Kim J, Doop ML, Gibson C, Lappin JS, Blake R, et al. (2006): Weakened center-surround interactions in visual motion processing in schizophrenia. Journal of Neuroscience. 26:11403–11412.

11. Butler PD, Zemon V, Schechter I, Saperstein AM, Hoptman MJ, Lim KO, et al. (2005): Early-stage visual processing and cortical amplification deficits in schizophrenia. Archives of General Psychiatry. 62:495–504.

12. Kéri S, Antal A, Szekeres G, Benedek G, Janka Z (2002): Spatiotemporal visual processing in schizophrenia. The Journal of Neuropsychiatry and Clinical Neurosciences. 14:190–196.

13. Slaghuis WA (1998): Contrast sensitivity for stationary and drifting spatial frequency gratings in positive- and negative-symptom schizophrenia. Journal of Abnormal Psychology. 107:49–62.

14. Yang E, Tadin D, Glasser DM, Hong SW, Blake R, Park S (2013): Visual context processing in schizophrenia. Clinical Psychological Science. 1:5–15.

15. Silverstein S, Bakshi S, Nuernberger S, Carpinello K, Wilkniss S (2005): Effects of stimulus structure and target-distracter similarity on the development of visual memory representations in schizophrenia. Cognitive Neuropsychiatry. 10:215–229.

16. Silverstein SM, Knight RA, Schwarzkopf SB, West LL, Osborn LM, Kamin D (1996): Stimulus configuration and context effects in perceptual organization in schizophrenia. Journal of Abnormal Psychology. 105:410–420.

17. Uhlhaas PJ, Phillips WA, Mitchell G, Silverstein SM (2006): Perceptual grouping in disorganized schizophrenia. Psychiatry Research. 145:105–117.

18. Uhlhaas PJ, Silverstein SM (2005): Perceptual organization in schizophrenia spectrum disorders: Empirical research and theoretical implications. 131:618.

19. Kéri S, Kelemen O, Benedek G, Janka Z (2004): Vernier threshold in patients with schizophrenia and in their unaffected siblings. Neuropsychology. 18:537–542.

20. O’Donnell BF, Swearer JM, Smith LT, Nestor PG, Shenton ME, McCarley RW (1996): Selective deficits in visual perception and recognition in schizophrenia. The American Journal of Psychiatry. 153:687–692.

21. Tek C, Gold J, Blaxton T, Wilk C, McMahon RP, Buchanan RW (2002): Visual perceptual and working memory impairments in schizophrenia. Archives of General Psychiatry. 59:146–153.

22. Butler PD, Harkavy-Friedman JM, Amador XF, Gorman JM (1996): Backward masking in schizophrenia: Relationship to medication status, neuropsychological functioning, and dopamine metabolism. Biological Psychiatry. 40:295–298.

23. Cadenhead KS, Serper Y, Braff DL (1998): Transient versus sustained visual channels in the visual backward masking deficits of schizophrenia patients. Biological Psychiatry. 43:132–138.

24. Green MF, Nuechterlein KH, Breitmeyer B, Mintz J (1999): Backward masking in unmedicated schizophrenic patients in psychotic remission: Possible reflection of aberrant cortical oscillation. American Journal of Psychiatry. 156:1367–1373.

25. Schechter I, Butler PD, Silipo G, Zemon V, Javitt DC (2003): Magnocellular and parvocellular contributions to backward masking dysfunction in schizophrenia. Schizophrenia Research. 64:91–101.

26. van de Ven V, Jagiela AR, Oertel-Knöchel V, Linden DE (2017): Reduced intrinsic visual cortical connectivity is associated with impaired perceptual closure in schizophrenia. NeuroImage: Clinical. 15:45–52.

27. Andrade GN, Butler JS, Peters GA, Molholm S, Foxe JJ (2016): Atypical visual and somatosensory adaptation in schizophrenia-spectrum disorders. Translational Psychiatry. 6:1–14.

28. Yeap S, Kelly SP, Sehatpour P, Magno E, Garavan H, Thakore JH, et al. (2008): Visual sensory processing deficits in Schizophrenia and their relationship to disease state. European Archives of Psychiatry and Clinical Neuroscience. 258:305–316.

29. Lalor EC, De Sanctis P, Krakowski MI, Foxe JJ (2012): Visual sensory processing deficits in schizophrenia: Is there anything to the magnocellular account? Schizophrenia Research. 139:246–252.

30. Yeap S, Kelly SP, Sehatpour P, Magno E, Javitt DC, Garavan H, et al. (2006): Early visual sensory deficits as endophenotypes for schizophrenia: High-density electrical mapping in clinically unaffected first-degree relatives. Archives of General Psychiatry. 63:1180–1188.

31. Sehatpour P, Dias EC, Butler PD, Revheim N, Guilfoyle DN, Foxe JJ, et al. (2010): Impaired Visual Object Processing Across an Occipital-Frontal-Hippocampal Brain Network in Schizophrenia: An Integrated Neuroimaging Study. Archives of General Psychiatry. 67:772–782.

32. González-Hernández JA, Pita-Alcorta C, Padrón A, Finalé A, Galán L, Martínez E, et al. (2014): Basic visual dysfunction allows classification of patients with schizophrenia with exceptional accuracy. Schizophrenia Research. 159:226–233.

33. Davenport ND, Sponheim SR, Stanwyck JJ (2006): Neural anomalies during visual search in schizophrenia patients and unaffected siblings of schizophrenia patients. Schizophrenia Research. 82:15–26.

34. Foxe JJ, Yeap S, Leavitt VM (2013): Brief monocular deprivation as an assay of short-term visual sensory plasticity in schizophrenia–”the binocular effect”. Frontiers in Psychiatry. 4:164.

35. Brenner CA, Lysaker PH, Wilt MA, O’Donnell BF (2002): Visual processing and neuropsychological function in schizophrenia and schizoaffective disorder. Psychiatry Research. 111:125–136.

36. Bruder G, Kayser J, Tenke C, Rabinowicz E, Friedman M, Amador X, et al. (1998): The time course of visuospatial processing deficits in schizophrenia: An event-related brain potential study. Journal of Abnormal Psychology. 107:399–411.

37. Foxe JJ, Murray MM, Javitt DC (2005): Filling-in in schizophrenia: a high-density electrical mapping and source-analysis investigation of illusory contour processing. Cerebral Cortex. 15:1914–1927.

38. Oribe N, Hirano Y, Kanba S, del Re EC, Seidman LJ, Mesholam-Gately R, et al. (2013): Early and late stages of visual processing in individuals in prodromal state and first episode schizophrenia: An ERP study. Schizophrenia Research. 146:95–102.

39. Uhlhaas PJ, Mishara AL (2007): Perceptual anomalies in schizophrenia: Integrating phenomenology and cognitive neuroscience. Schizophrenia Bulletin. 33:142–156.

40. Sergi MJ, Green MF (2003): Social perception and early visual processing in schizophrenia. Schizophrenia Research. 59:233–241.

41. Rassovsky Y, Horan WP, Lee J, Sergi MJ, Green MF (2011): Pathways between early visual processing and functional outcome in schizophrenia. Psychological Medicine. 41:487–497.

42. Green MF, Kern RS, Braff DL, Mintz J (2000): Neurocognitive deficits and functional outcome in schizophrenia: are we measuring the “right stuff”? Schizophrenia Bulletin. 26:119–136.

43. Crumlish N, Whitty P, Clarke M, Browne S, Kamali M, Gervin M, et al. (2009): Beyond the critical period: longitudinal study of 8-year outcome in first-episode non-affective psychosis. The British Journal of Psychiatry. 194:18–24.

44. Birchwood M, Todd P, Jackson C (1998): Early intervention in psychosis: The critical period hypothesis. The British Journal of Psychiatry. 172:53–59.

45. Eack SM, Greenwald DP, Hogarty SS, Keshavan MS (2010): One-year durability of the effects of cognitive enhancement therapy on functional outcome in early schizophrenia. Schizophrenia Research. 120:210–216.

46. ten Velden Hegelstad W, Haahr U, Larsen TK, Auestad B, Barder H, Evensen J, et al. (2013): Early detection, early symptom progression and symptomatic remission after ten years in a first episode of psychosis study. Schizophrenia Research. 143:337–343.

47. Sklar AL, Coffman BA, Haas G, Ghuman A, Cho R, Salisbury DF (2020): Inefficient visual search strategies in the first-episode schizophrenia spectrum. Schizophrenia Research. 224:126–132.

48. Sklar AL, Coffman BA, Salisbury DF (2020): Localization of early-stage visual processing deficits at schizophrenia spectrum illness onset using magnetoencephalography. Schizophrenia Bulletin. 46:955–963.

49. Sklar AL, Coffman BA, Salisbury DF (2021): Fronto-parietal network function during cued visual search in the first-episode schizophrenia spectrum. Journal of Psychiatric Research 141:339–345.

50. Haenschel C, Bittner RA, Haertling F, Rotarska-Jagiela A, Maurer K, Singer W, et al. (2007): Contribution of impaired early-stage visual processing to working memory dysfunction in adolescents with schizophrenia: a study with event-related potentials and functional magnetic resonance imaging. Archives of General Psychiatry. 64:1229–1240.

51. Yeap S, Kelly SP, Thakore JH, Foxe JJ (2008): Visual sensory processing deficits in first-episode patients with Schizophrenia. Schizophrenia Research. 1:340–343.

52. McCabe KL, Marlin S, Cooper G, Morris R, Schall U, Murphy DG, et al. (2016): Visual perception and processing in children with 22q11. 2 deletion syndrome: associations with social cognition measures of face identity and emotion recognition. Journal of Neurodevelopmental Disorders. 8:1–8.

53. Biria M, Tomescu MI, Custo A, Cantonas LM, Song KW, Schneider M, et al. (2018): Visual processing deficits in 22q11. 2 deletion syndrome. NeuroImage: Clinical. 17:976–986.

54. Magnée MJ, Lamme VA, de Sain-van der Velden MG, Vorstman JA, Kemner C (2011): Proline and COMT status affect visual connectivity in children with 22q11. 2 deletion syndrome. PLoS One. 6:e25882.

55. Francisco AA, Foxe JJ, Horsthuis DJ, DeMaio D, Molholm S (2020): Assessing auditory processing endophenotypes associated with Schizophrenia in individuals with 22q11. 2 deletion syndrome. Translational Psychiatry. 10:1–11.

56. Francisco AA, Horsthuis DJ, Popiel M, Foxe JJ, Molholm S (2020): Atypical response inhibition and error processing in 22q11. 2 Deletion Syndrome and schizophrenia: Towards neuromarkers of disease progression and risk. NeuroImage: Clinical. 27:102351.

57. Andrade GN, Butler JS, Mercier MR, Molholm S, Foxe JJ (2015): Spatioltemporal dynamics of adaptation in the human visual system: A highldensity electrical mapping study. European Journal of Neuroscience. 41:925–939.

58. Dias EC, Butler PD, Hoptman MJ, Javitt DC (2011): Early sensory contributions to contextual encoding deficits in schizophrenia. Archives of General Psychiatry. 68:654–664.

59. Butler PD, Martinez A, Foxe JJ, Kim D, Zemon V, Silipo G, et al. (2007): Subcortical visual dysfunction in schizophrenia drives secondary cortical impairments. Brain. 130:417–430.

60. Pinheiro AP, Liu T, Nestor PG, McCarley RW, Gonçalves OF, Niznikiewicz MA (2013): Visual emotional information processing in male schizophrenia patients: Combining ERP, clinical and behavioral evidence. Neuroscience Letters. 550:75–80.

61. Pokorny VJ, Lano TJ, Schallmo MP, Olman CA, Sponheim SR (2021): Reduced influence of perceptual context in schizophrenia: Behavioral and neurophysiological evidence. Psychological Medicine. 51:786–794.

62. Doniger GM, Foxe JJ, Murray MM, Higgins BA, Javitt DC (2002): Impaired visual object recognition and dorsal/ventral stream interaction in schizophrenia. Archives of General Psychiatry. 59:1011–1020.

63. Foxe JJ, Doniger GM, Javitt DC (2001): Early visual processing deficits in schizophrenia: Impaired P1 generation revealed by high-density electrical mapping. Neuroreport. 12:3815–3820.

64. Schaer M, Glaser B, Ottet MC, Schneider M, Cuadra MB, Debbané M, et al. (2010): Regional cortical volumes and congenital heart disease: a MRI study in 22q11. 2 deletion syndrome. Journal of Neurodevelopmental Disorders. 2:224–234.

65. Scariati E, Schaer M, Richiardi J, Schneider M, Debbané M, Van De Ville D, et al. (2014): Identifying 22q11. 2 deletion syndrome and psychosis using resting-state connectivity patterns. Brain Topography. 27:808–821.

66. Cattan S, Bachatene L, Bharmauria V, Jeyabalaratnam J, Milleret C, Molotchnikoff S (2014): Comparative analysis of orientation maps in areas 17 and 18 of the cat primary visual cortex following adaptation. European Journal of Neuroscience. 40:2554–2563.

67. Muller JR, Metha AB, Krauskopf J, Lennie P (1999): Rapid adaption in visual cortex to the structure of images. Science. 285:1405.

68. Wissig SC, Kohn A (2012): The influence of surround suppression on adaptation effects in primary visual cortex. Journal of Neurophysiology. 107:3370–3384.

69. Wark B, Lundstrom BN, Fairhall A (2007): Sensory Adaptation. Current Opinion in Neurobiology. 17:423–429.

70. Näätänen R, Picton T (1987): The N1 wave of the human electric and magnetic response to sound: A review and an analysis of the component structure. Psychophysiology. 24:375–425.

71. Roth WT, Krainz PL, Ford JM, Tinklenberg JR, Rothbart RM, Kopell BS (1976): Parameters of temporal recovery of the human auditory evoked potential. Electroencephalography and Clinical Neurophysiology. 40:623–632.

72. Budd TW, Barry RJ, Gordon E, Rennie C, Michie PT (1998): Decrement of the N1 auditory event-related potential with stimulus repetition: Habituation vs. refractoriness. International Journal of Psychophysiology. 31:51–68.

73. Muller-Gass A, Marcoux A, Jamshidi P, Campbell K (2008): The effects of very slow rates of stimulus presentation on event-related potential estimates of hearing threshold. International Journal of Audiology. 47:34–43.

74. Pereira DR, Cardoso S, Ferreira-Santos F, Fernandes C, Cunha-Reis C, Paiva TO, et al. (2014): Effects of inter-stimulus interval (ISI) duration on the N1 and P2 components of the auditory event-related potential. International Journal of Psychophysiology. 94:311–318.

75. Rosburg T, Zimmerer K, Huonker R (2010): Short-term habituation of auditory evoked potential and neuromagnetic field components in dependence of the interstimulus interval. Experimental Brain Research. 205:559–570.

76. Larsson J, Smith AT (2012): fMRI repetition suppression: neuronal adaptation or stimulus expectation? Cerebral Cortex. 22:567–576.

77. Wastell DG, Kleinman D (1980): A psychoanatomical investigation of the locus of the mechanism responsible for the refractoriness of the visual vertex potential. Perception & Psychophysics. 27:149–152.

78. Gjini K, Sundaresan K, Boutros NN (2008): Electroencephalographic evidence of sensory gating in the occipital visual cortex. Neuroreport. 19:1519–1522.

79. Adler LE, Waldo MC, Freedman R (1985): Neurophysiologic studies of sensory gating in schizophrenia: comparison of auditory and visual responses. Biological Psychiatry. 20:1284–1296.

80. Wechsler D (2008): WAIS-IV Administration and Scoring Manual. San Antonio, TX: Psychological Corporation.

81. Wechsler D (2014): WISC-V: Technical and Interpretive Manual. Bloomington, MN: Pearson.

82. First MB, Williams JBW, Karg RS, Spitzer RL (2015): Structured Clinical Interview for DSM-5— Research Version (SCID-5 for DSM-5, research version; SCID-5-RV). Arlington, VA: American Psychiatric Association.

83. Hien D, Matzner FJ, First MB, Spitzer RL, Gibbon M, Williams JBW (1994): Structured Clinical Interview for DSM-IV-Child Edition (Version 1.0). New York: Columbia University.

84. Delorme A, Makeig S (2004): EEGLAB: an open source toolbox for analysis of single-trial EEG dynamics including independent component analysis. Journal of Neuroscience Methods. 134:9–21.

85. Horsthuis DJ, Francisco AA (2021): EEG_to_ERP_pipeline_stats_R (V1.1). Zenodo: https://doi.org/10.5281/zenodo.5590389.

86. Bates D, Mächler M, Bolker BM, Walker SC (2014): Ime4: Linear mixed-effects models using Eigen and S4 (R package Version 1.1-7) [Computer software]. Retrieved from http://CRANR-projectorg/package=lme4.

87. RCoreTeam (2014): R: A language and environment for statistical computing 3.3.2 ed. Vienna, Austria: R Foundation for Statistical Computing.

88. Holm S (1979): A simple sequentially rejective multiple test procedure. Scandinavian Journal of Statistics. 6:65–70.

89. Cantonas LM, Tomescu MI, Biria M, Jan RK, Schneider M, Eliez S, et al. (2019): Abnormal development of early auditory processing in 22q11.2 Deletion Syndrome. Translational Psychiatry. 9:1–12.

90. Rihs TA, Tomescu MI, Britz J, Rochas V, Custo A, Schneider M, et al. (2013): Altered auditory processing in frontal and left temporal cortex in 22q11.2 deletion syndrome: A group at high genetic risk for schizophrenia. Psychiatry Research. 212:141–149.

91. Didriksen M, Fejgin K, Nilsson SRO, Birknow MR, Grayton HM, Larsen PH, et al. (2017): Persistent gating deficit and increased sensitivity to NMDA receptor antagonism after puberty in a new mouse model of the human 22q11.2 microdeletion syndrome: a study in male mice. Journal of Psychiatry & Neuroscience. 42:48–58.

92. Lalor EC, Yeap S, Reilly RB, Pearlmutter BA, Foxe JJ (2008): Dissecting the cellular contributions to early visual sensory processing deficits in schizophrenia using the VESPA evoked response. Schizophrenia Research. 98:256–264.

93. Butler PD, Schechter I, Zemon V, Schwartz SG, Greenstein VC, Gordon J, et al. (2001): Dysfunction of early-stage visual processing in schizophrenia. American Journal of Psychiatry. 158:1126–1133.

94. Knebel JF, Javitt DC, Murray MM (2011): Impaired early visual response modulations to spatial information in chronic schizophrenia. Psychiatry Research: Neuroimaging. 193:168–176.

95. Johnson SC, Lowery N, Kohler C, Turetsky BI (2005): Global–local visual processing in schizophrenia: evidence for an early visual processing deficit. Biological Psychiatry. 58:937–946.

96. Plomp G, Roinishvili M, Chkonia E, Kapanadze G, Kereselidze M, Brand A, et al. (2013): Electrophysiological evidence for ventral stream deficits in schizophrenia patients. Schizophrenia Bulletin. 39:547–554.

97. Yeap S, Kelly SP, Reilly RB, Thakore JH, Foxe JJ (2009): Visual sensory processing deficits in patients with bipolar disorder revealed through high-density electrical mapping. Journal of Psychiatry & Neuroscience. 34:459–464.

98. Oranje B, van Berckel BNM, Kemner C, van Ree JM, Kahn RS, Verbaten MN (2000): The effects of a sub-anaesthetic dose of ketamine on human selective attention. Neuropsychopharmacology. 22:293–302.

99. Umbricht D, Schmid L, Koller R, Vollenweider FX, Hell D, Javitt DC (2000): Ketamine-induced deficits in auditory and visual context-dependent processing in healthy volunteers. Archives of General Psychiatry. 57:1139–1147.

100. Schwertner A, Zortea M, Torres FV, Caumo W (2018): Effects of subanesthetic ketamine administration on visual and auditory event-related potentials (ERP) in humans: A systematic review. Frontiers in Behavioral Neuroscience. 12:70.

101. Vann Jones S, Banerjee S, Smith AD, Refsum H, Lennox B (2017): Elevated homocysteine and N-methyl-D-aspartate-receptor antibodies as a cause of behavioural and cognitive decline in 22q11. 2 deletion syndrome. Oxford Medical Case Reports. 2017:omx076.

102. Coyle JT (2012): NMDA receptor and schizophrenia: A brief history. Schizophrenia Bulletin. 38:920–926.

103. Donohoe G, Morris DW, De Sanctis P, Magno E, Montesi JL, Garavan HP, et al. (2008): Early visual processing deficits in dysbindin-associated schizophrenia. Biological Psychiatry. 63:484–489.

104. O’Donoghue T, Morris DW, Fahey C, Costa AD, Foxe JJ, Hoerold D, et al. (2012): A NOS1 variant implicated in cognitive performance influences evoked neural responses during a high density EEG study of early visual perception. Human Brain Mapping. 33:1202–1211.

105. Zinkstok J, Schmitz N, Van Amelsvoort T, Moeton M, Baas F, Linszen D (2008): Genetic variation in COMT and PRODH is associated with brain anatomy in patients with schizophrenia. Genes, Brain and Behavior. 7:61–69.

106. Chen LC, Stropahl M, Schönwiesner M, Debener S (2017): Enhanced visual adaptation in cochlear implant users revealed by concurrent EEG-fNIRS. NeuroImage. 146:600–608.

107. Bradley C, Joyce N, Garcia-Larrea L (2016): Adaptation in human somatosensory cortex as a model of sensory memory construction: A study using high-density EEG. Brain Structure and Function. 221:421–431.

108. Jääskeläinen IP, Ahveninen J, Andermann ML, Belliveau JW, Raij T, Sams M (2011): Short-term plasticity as a neural mechanism supporting memory and attentional functions. Brain Research. 1422:66–81.

109. Simon TJ (2008): A new account of the neurocognitive foundations of impairments in space, time, and number processing in children with chromosome 22q11. 2 deletion syndrome. Developmental Disabilities Research Reviews. 14:52–58.

110. Antshel KM, Fremont W, Kates WR (2008): The neurocognitive phenotype in velolcardiolfacial syndrome: A developmental perspective. Developmental Disabilities Research Reviews. 14:43–51.

111. Bearden CE, Woodin MF, Wang PP, Moss E, McDonald-McGinn D, Zackai E, et al. (2001): The neurocognitive phenotype of the 22q11. 2 deletion syndrome: selective deficit in visual-spatial memory. Journal of Clinical and Experimental Neuropsychology. 23:447–464.

112. Shapiro HM, Takarae Y, Harvey DJ, Cabaral MH, Simon TJ (2012): A cross-sectional study of the development of volitional control of spatial attention in children with chromosome 22q11. 2 deletion syndrome. Journal of Neurodevelopmental Disorders. 4:1–12.

113. Campbell LE, Azuma R, Ambery F, Stevens A, Smith A, Morris RG, et al. (2010): Executive functions and memory abilities in children with 22q11. 2 deletion syndrome. Australian & New Zealand Journal of Psychiatry. 44:364–371.

114. Palermo A, Giglia G, Vigneri S, Cosentino G, Fierro B, Brighina F (2011): Does habituation depend on cortical inhibition? Results of a rTMS study in healthy subjects. Experimental Brain Research. 212:101–107.

115. Kohn A (2007): Visual adaptation: Physiology, mechanisms, and functional benefits. Journal of Neurophysiology. 97:3155–3164.

116. Kapur S (2003): Psychosis as a state of aberrant salience: A framework linking biology, phenomenology, and pharmacology in schizophrenia. American Journal of Psychiatry. 160:13–23.

117. Silverstein SM, Paterno D, Cherneski L, Green S (2018): Optical coherence tomography indices of structural retinal pathology in schizophrenia. Psychological Medicine. 48:2023–2033.

118. Butler PD, Silverstein SM, Dakin SC (2008): Visual perception and its impairment in schizophrenia. Biological Psychiatry. 64:40–47.

119. Buch ER, Young S, Contreras-Vidal JL (2003): Visuomotor adaptation in normal aging. Learning & Memory. 10:55–63.

120. Braff DL, Saccuzzo DP (1982): Effect of antipsychotic medication on speed of information processing in schizophrenic patients. American Journal of Psychiatry. 139:1127–1130.

121. Harvey PD, Docherty NM, Serper MR, Rasmussen M (1990): Cognitive deficits and thought disorder: II. An 8-month followup study. Schizophrenia Bulletin. 16:147–156.

122. Cadenhead KS, Geyer MA, Butler RW, Perry W, Sprock J, Braff DL (1997): Information processing deficits of schizophrenia patients: Relationship to clinical ratings, gender and medication status. Schizophrenia Research. 28:51–62.

123. Gjini K, Burroughs S, Boutros NN (2011): Relevance of attention in auditory sensory gating paradigms in schizophrenia: A pilot study. Journal of Psychophysiology. 25:60–66.

124. Rosburg T, Trautner P, Elger CE, Kurthen M (2009): Attention effects on sensory gating–intracranial and scalp recordings. NeuroImage. 48:554–563.

